# New-onset infective endocarditis in diabetic patients receiving SGLT2I, DPP4I and GLP1a: A population-based cohort study

**DOI:** 10.1101/2023.08.06.23293730

**Authors:** Oscar Hou-In Chou, Tianyu Gao, Cheuk To Chung, Fengshi Jing, Jeffrey Shi Kai Chan, Danish Iltaf Satti, Ronald TK Pang, Sharen Lee, Wing Tak Wong, Tong Liu, Gregory Y H Lip, Bernard Man Yung Cheung, Gary Tse, Jiandong Zhou

**Author notes:** Contributed equally (Oscar Hou-In Chou, Tianyu Gao). Correspondence to: Gary Tse, MD, Ph.D., FRCP, FFPH Kent and Medway Medical School, Canterbury, United Kingdom Tianjin Institute of Cardiology, The Second Hospital of Tianjin Medical University, Tianjin 300211, China School of Nursing and Health Studies, Hong Kong Metropolitan University, Hong Kong, China, Jiandong Zhou, Ph.D. Division of Health Science, Warwick Medical School, University of Warwick, Coventry, United Kingdom.

## Abstract

**Background:** Sodium-glucose cotransporter-2 inhibitors (SGLT2I) have been suggested to have beneficial effects against infection. However, the comparative risks of new onset infective endocarditis between SGLT2Is, dipeptidyl peptidase-4 inhibitors (DPP4Is) and glucagon-like peptide-1 receptor agonist (GLP1a) remain unknown.

**Objective:** This real-world study aims to compare the risks of infective endocarditis upon exposure to SGLT2I and dipeptidyl peptidase-4 inhibitors (DPP4I).

**Methods:** This was a retrospective population-based cohort study of patients with type-2 diabetes mellitus (T2DM) on either SGLT2I or DPP4I between 1st January 2015 and 31st December 2020 using a territory-wide registry in Hong Kong. The primary outcome was new-onset infective endocarditis. The secondary outcome was cardiovascular-related mortality. Propensity score matching (1:1 ratio) using the nearest neighbour search was performed. Multivariable Cox regression was applied to identify significant associations. A three-arm sensitivity analysis including the GLP1a cohort was conducted.

**Results:** This cohort included 75638 T2DM patients (median age: 62.3 years old [SD: 12.8]; 55.79 % males). The SGLT2I and DPP4I groups consisted of 28774 patients and 46864 patients, respectively. After matching, 104 and 161 infective endocarditis in the SGLT2I and DPP4I groups occurred over a follow-up of 5.6 years. SGLT2I use was associated with lower risks of infective endocarditis (Hazard ratio [HR]: 0.58; 95% Confidence Interval [CI]: 0.41-0.81) and cardiovascular mortality (HR: 0.49; 95% CI: 0.33-0.72) compared to DPP4I use after adjustments for demographics, comorbidities, medications, renal function, and HbA1c levels. Similar associations were observed in subgroup analyses regardless of gender, hypertension, prior valvular disease, renal disease, or immunodeficiency. In the sensitivity analysis, SGLT2I was not associated with lower risks of infective endocarditis compared to GLP1a. The results remained consistent in the competing risk and the other sensitivity analyses.

**Conclusions:** SGLT2I use was associated with lower risks of new-onset infective endocarditis compared to DPP4I after adjustments.

**Illustrated abstract:** 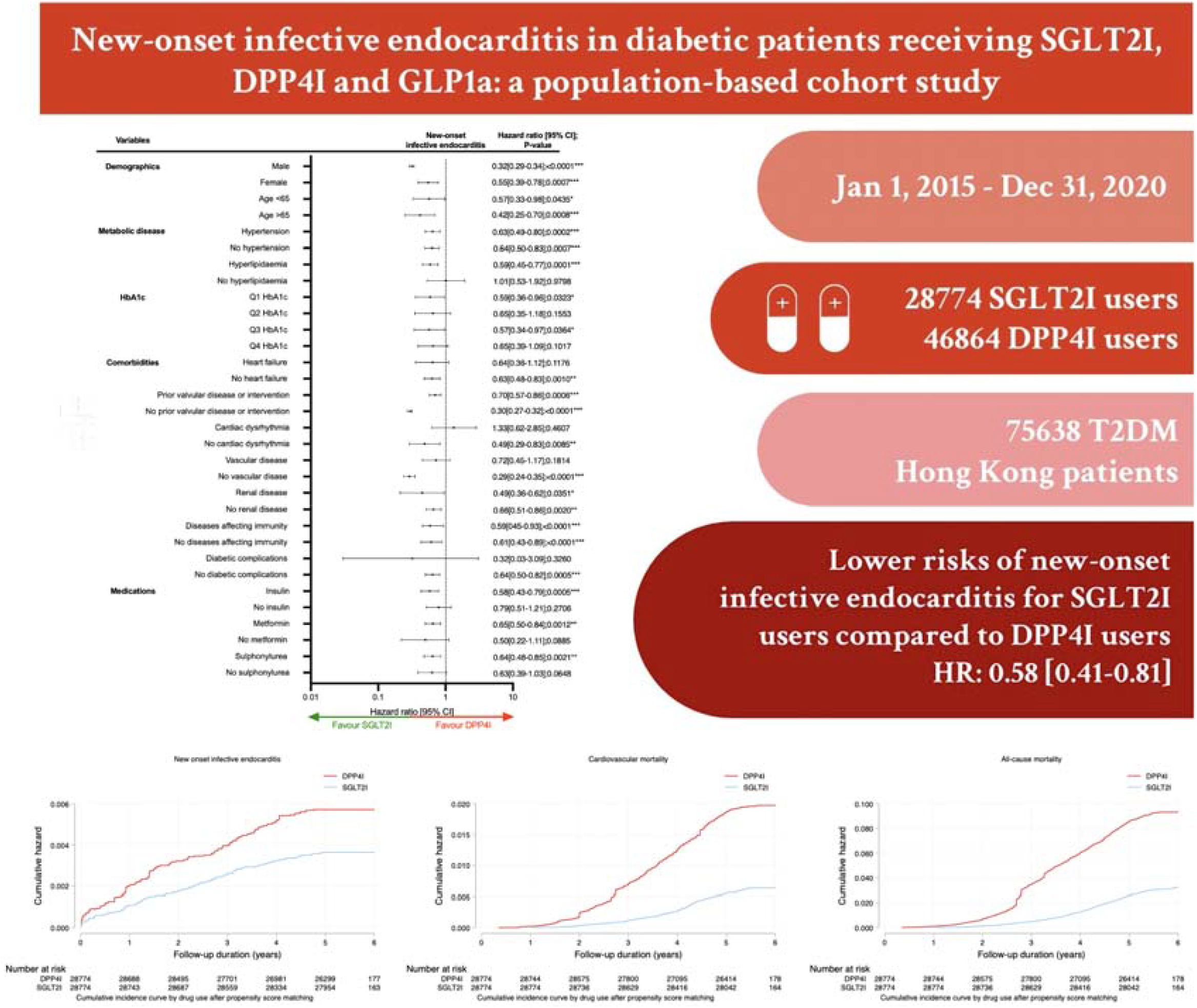

## Introduction

The prevalence of type 2 diabetes mellitus (T2DM) remains a significant burden on the global healthcare system. It is well established that diabetic patients are at higher risk of various types of infections,^1^ including respiratory infections^2^ and cardiac infections.^3^ Patients with diabetes are at increased risk of infective endocarditis, related complications, and adverse events compared to patients without diabetes. ^4-6^ Subsequently, this instigated interest in further understanding the potential influence of oral hypoglycaemic agents against infective endocarditis. While a plethora of studies show that novel agents such as SGLT2I and DPP4I can effectively improve glycaemic parameters and exert cardiovascular benefits,^7, 8^ their influence on cardiac infections remain ambiguous.

Previous studies have demonstrated that a drastic difference in the prevalence of IE by *Staphylococcus sp.* between diabetic patients who were insulin users compared to those who used oral hypoglycaemic drugs or were non-diabetic.^9^ Novel anti-diabetic medications, such as sodium-glucose cotransporter 2 (SGLT2) inhibitors, dipeptidyl peptidase-4 (DPP4) inhibitors, and glucagon-like peptide-1 receptor agonist (GLP1a) have shown promising results in reducing various types of infections.^10, 11^ Resultantly, studies have postulated that the anti-infective effects of these agents may potentially extend to reducing the prevalence of heart infections^12^. To the best of our knowledge, there is little to no evidence of large cohort studies investigating the association between novel anti-diabetic medications and the prevalence of infective endocarditis.

Thereby, this study aims to examine the role of SGLT2I and DPP4I with new-onset infective endocarditis in a cohort of T2DM patients from Hong Kong.

## Methods

### Study population

This study was approved by the Institutional Review Board of the University of Hong Kong/Hospital Authority Hong Kong West Cluster (HKU/HA HKWC IRB) (UW-20-250) and complied with the Declaration of Helsinki. This was a retrospective population-based study of prospectively collected electronic health records using the Clinical Data Analysis and Reporting System (CDARS) by the Hospital Authority (HA) of Hong Kong. The records cover both public hospitals and private clinics in Hong Kong. It was verified that over 90% of the identified T2DM patients were under the HA.^13, 14^ This system has been used extensively by our teams and other research teams in Hong Kong. The system contains data on disease diagnosis, laboratory results, past comorbidities, clinical characteristics, and medication prescriptions. The system has been used by our Hong Kong research team to perform comparative studies.^15, 16^ T2DM patients who were administered with SGLT2I or DPP4I in centres under the Hong Kong HA, between 1^st^ January 2015, to 31^st^ December 2020, were included. The glucagon-like peptide-1 receptor agonist (GLP1a) cohort comprised of patients on GLP1a between 1^st^ January 2015, to 31^st^ December 2020 was included for sensitivity analysis to demonstrate the relative effects amongst the second line oral anti-diabetic agents.

### Predictors and variables

Patients’ demographics include gender and age of initial drug use (baseline), clinical and biochemical data were extracted for the present study. Prior comorbidities were extracted in accordance with the *International Classification of Diseases Ninth Edition* (ICD-9) codes (**Supplementary Table 1**). The diabetes duration was calculated by examining the earliest date amongst the first date of (1) diagnosis using of ICD-9; (2) Hba1c >=6.5%; (3) Fasting glucose >= 7.0 mmol/l or Random glucose 11.1 mmol/l; (4) using anti-diabetic medications. The patients on financial aid were defined by patients on Comprehensive Social Security Assistance (CSSA) scheme, higher disability allowance, normal disability allowance, wavier and other financial aids in Hong Kong. The number of hospitalisations in the year prior the index days were extracted. The Charlson’s standard comorbidity index was calculated.^17^ The duration and frequency of SGLT2I and DPP4I usage were calculated. The baseline laboratory examinations, including the glucose profiles and renal function tests were extracted. The microbiology testing results were also extracted to identify the potential causative agents for infective endocarditis. The estimated glomerular filtration rate (eGFR) was calculated using the abbreviated modification of diet in renal disease (MDRD) formula.^18^ The following patient groups were excluded: those who (1) with prior infective endocarditis; (2) with congenital heart disease; (3) patients who died within 30 days after initial drug exposure; (4) without complete demographics; (5) under 18 years old **(Figure 1)**. To account for the incomplete laboratory data, the multiple imputation by chained equations was performed according to the previous published study ^19^.

**Figure 1.**
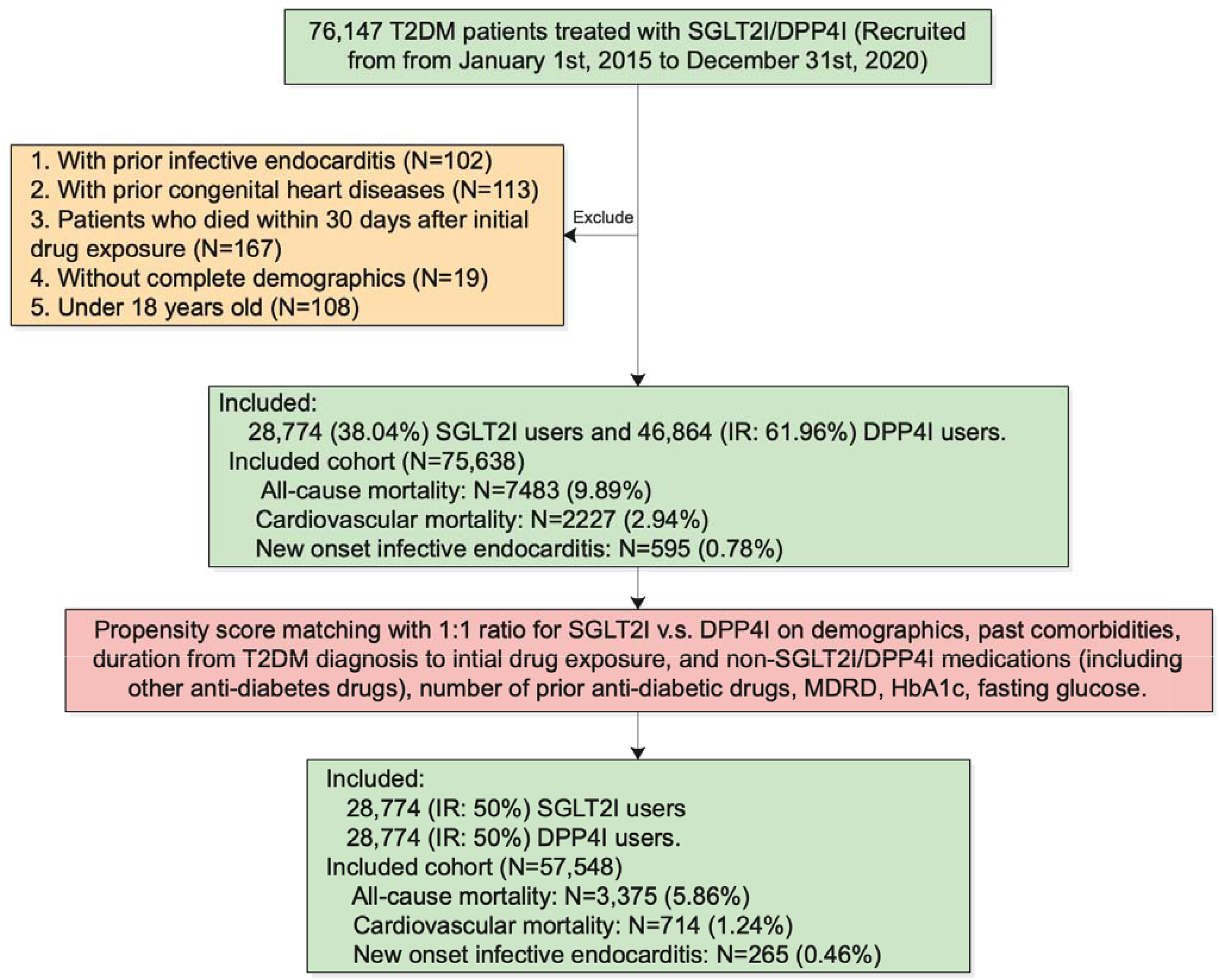
Procedures of data processing for the study cohort. SGLT2I: Sodium-glucose cotransporter-2 inhibitors; DPP4I: Dipeptidyl peptidase-4 inhibitors. MDRD: modification of diet in renal disease.

### Study outcomes

The primary outcome of this study was new-onset infective endocarditis defined using either ICD-9 code or 2 separate positive blood culture results according to the modified Duke’s criteria (**Supplementary Table 1**) after the index date of the drug use, according to the verified published literature.^20^ The secondary outcomes were cancer-related mortality and all-cause mortality. Mortality data were obtained from the Hong Kong Death Registry, a population-based official government registry with the registered death records of all Hong Kong citizens linked to CDARS. Mortality was recorded using the *International Classification of Diseases Tenth Edition* (ICD-10) coding. The as-treat approach was adopted which patients were censored at treatment discontinuation or switching of the comparison medications. The endpoint date of interest for eligible patients was the event presentation date. The endpoint for those without primary outcome was the mortality date or the end of the study period (31^st^ December 2020).

### Statistical analysis

Descriptive statistics are used to summarize baseline clinical and biochemical characteristics of patients with SGLT2I and DPP4I use. For baseline clinical characteristics, continuous variables were presented as mean (95% confidence interval/standard deviation) and the categorical variables were presented as total numbers (percentage). Propensity score matching generated by logistic regression with 1:1 ratio for SGLT2I use versus DPP4I use based on demographics, non-SGLT2I/DPP4I medications, number of prior anti-diabetic drugs, prior comorbidities, renal function, duration from T2DM diagnosis initial drug exposure, HbA1c and fasting glucose were performed using the nearest neighbour search strategy with a calliper of 0.1. Propensity score matching was performed using Stata software (Version 16.0). Baseline characteristics between patients with SGLT2I and DPP4I use before and after matching were compared with absolute standardized mean difference (SMD), with SMD<0.10 regarded as well-balanced between the two groups.

The cumulative incidence curves for the primary outcomes and secondary outcomes were constructed. Proportional Cox regression models were used to identify significant risk predictors of adverse study outcomes, with adjustment for demographics, comorbidities, number of prior hospitalisations, medication profile, renal function, glycaemic tests, and the duration of T2DM. The log-log plot was used to verify the proportionality assumption for the proportional Cox regression models. Subgroup analysis was conducted to confirm the association amongst patients with different clinical important predictors accounting to the diabetic and the metabolic profile, as well as the comorbidities and medications associated with infective endocarditis.

Cause-specific and sub-distribution hazard models were conducted to consider possible competing risks. Multiple propensity adjustment approaches were used, including propensity score stratification ^21^, propensity score matching with inverse probability of treatment weighting ^22^ and propensity score matching with stable inverse probability weighting ^23^. The three arm sensitivity results involving glucagon-like peptide-1 receptor agonist (GLP1a) using stabilized IPTW were conducted to test the association and choice amongst the novel second-line anti-diabetic medications. Sensitivity analysis results with consideration of one-year lag time effects was conducted. Patients with CKD stage 4/5 (eGFR <30 mL/min/1.73m^2), peritoneal dialysis or haemodialysis who may be contraindicated with SGLT2I were excluded in the sensitivity analysis. Furthermore, patients with prior drug abuse, or with financial aids were excluded while the patients who developed infective endocarditis within 30 days after medications initiation were included in the sensitivity analysis. Lastly, a sensitivity analysis was included by defining new-onset infective endocarditis as including only patients with 2 separate blood cultures positive with organisms typical of infective endocarditis according to the culture criteria in the modified Duke’s criteria.

## Results

### Basic characteristics

This territory-wide cohort included 76147 T2DM patients treated with SGLT2I or DPP4I between 1st January 2015 and 31st December 2020 in Hong Kong. The patients were followed up until 31st December 2020, switching treatment, or until their deaths **(Figure 1)**. After exclusion, this cohort included 75638 patients, in which there were 28774 SGLT2I users and 46864 DPP4I users.

The DPP4I and SGLT2I users were comparable after propensity score matching with nearest neighbour search strategy with calliper of 0.1, and the proportional hazard assumption was tested **(Supplementary Figure 1)**. After the propensity score matching, the baseline characteristic of the SGLT2I and the DPP4I users were well-balanced, except baseline age (SGLT2I: 57.9 vs DPP4I: 59.2; SMD = 0.12), other lipid lowering drugs (SGLT2I: 35.20% vs DPP4I: 41.46%; SMD = 0.13), abbreviated MDRD (SGLT2I: 89.8 mL/min/1.73m^2 vs DPP4I: 88.2 mL/min/1.73m^2; SMD = 0.13). In the matched cohort, 265 patients developed new onset infective endocarditis. Amongst the infective endocarditis patients, 102 (38.4%) of the patients had the pathogen data. The most common infection identified was *Staphylococcus aureus* (41.2%), followed by *Streptococcus viridans* (35.3%), other microorganisms (6.86%), other *Streptococcus spp.* (5.88%), and the HACEK group bacteria (4.90%) **(Supplementary Figure 2)**. The characteristics of patients are shown in **Table 1**.

**Table 1.**
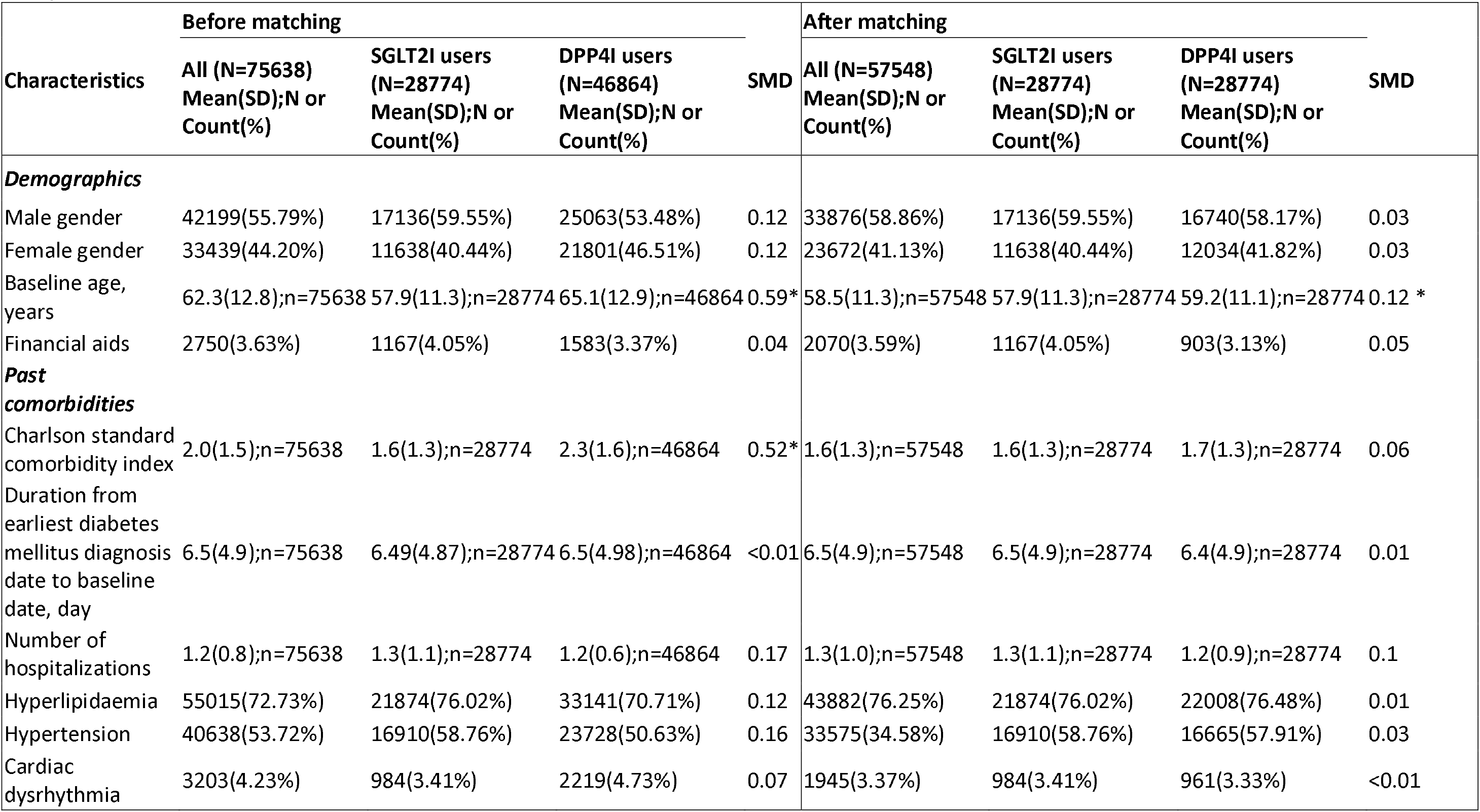
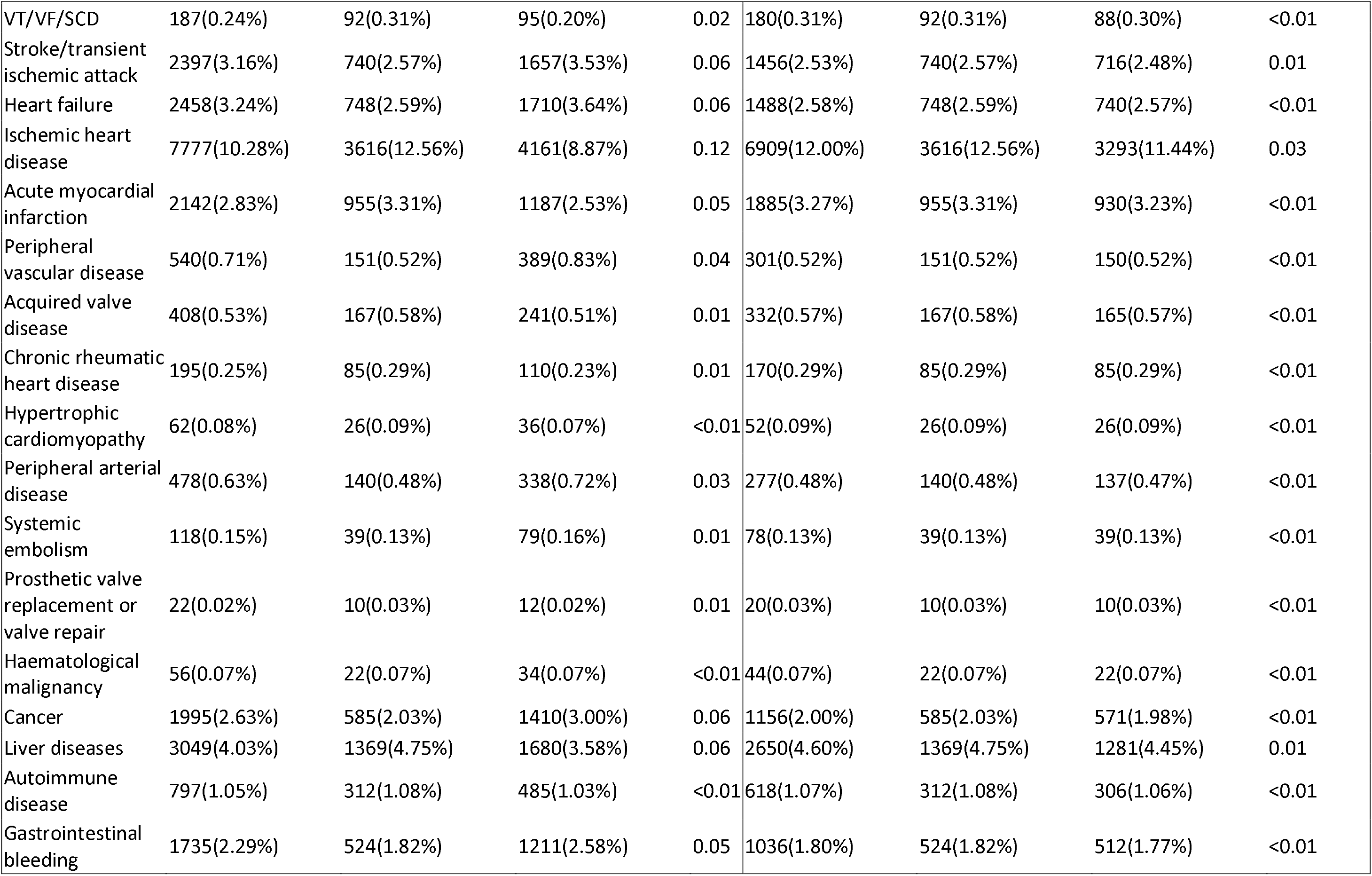
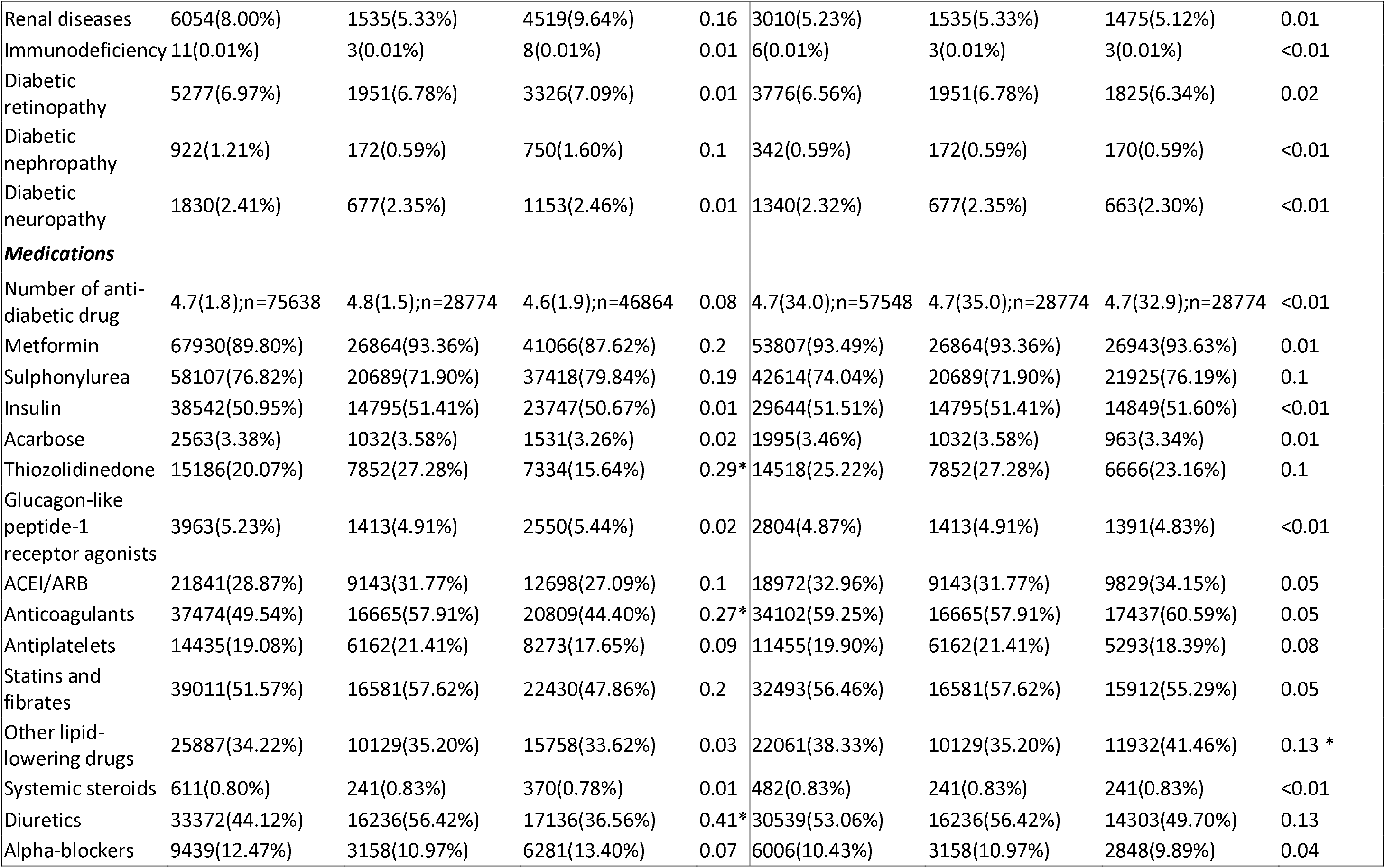
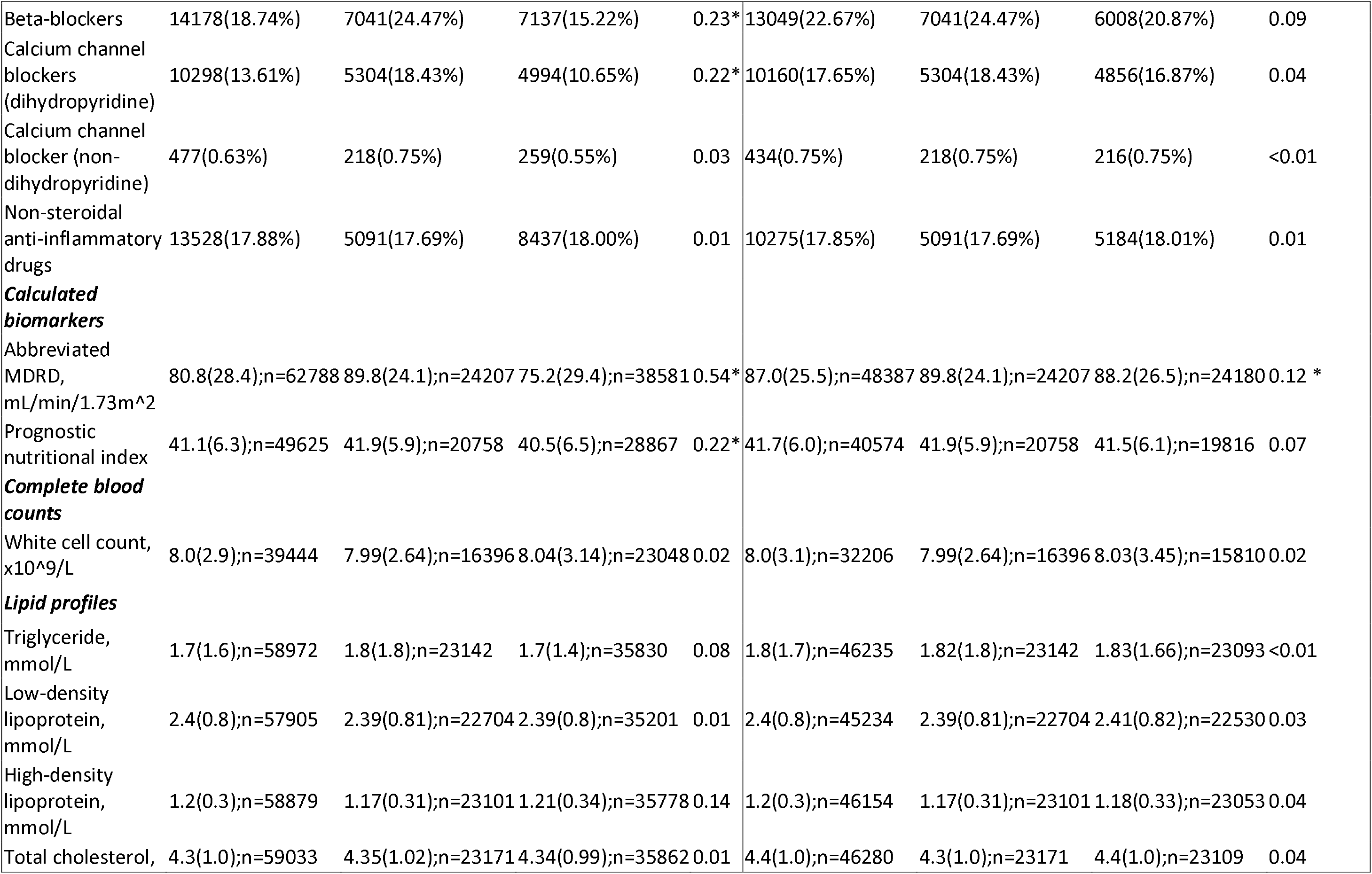
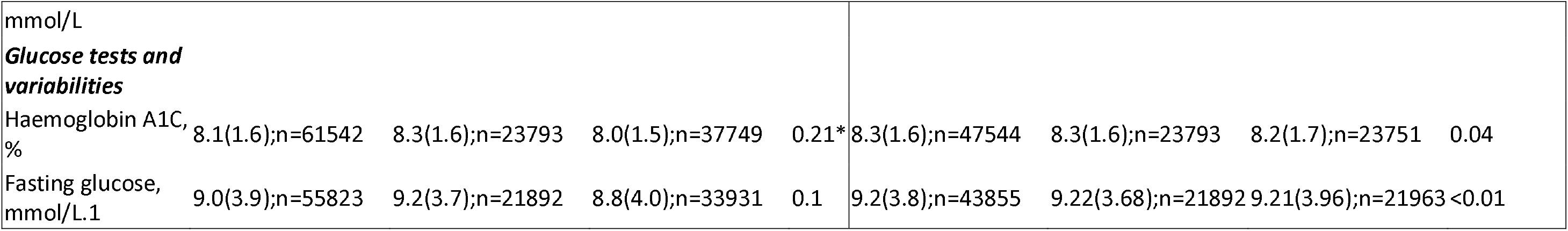
Baseline and clinical characteristics of patients with SGLT2I v.s. DPP4I use before and after propensity score matching (1:1). * for SMD>0.1; SGLT2I: sodium glucose cotransporter-2 inhibitor; DPP4I: dipeptidyl peptidase-4 inhibitor; SD: standard deviation; CV: Coefficient of variation; MDRD: modification of diet in renal disease; ACEI: angiotensin-converting enzyme inhibitors; ARB: angiotensin II receptor blockers.

| Characteristics | Before matching |  |  |  | After matching |  |  |  |
| --- | --- | --- | --- | --- | --- | --- | --- | --- |
|  | All (N=75638)<br>Mean(SD);N or<br>Count(%) | SGLT2I users<br>(N=28774)<br>Mean(SD);N or<br>Count(%) | DPP4I users<br>(N=46864)<br>Mean(SD);N or<br>Count(%) | SMD | All (N=57548)<br>Mean(SD);N or<br>Count(%) | SGLT2I users<br>(N=28774)<br>Mean(SD);N or<br>Count(%) | DPP4I users<br>(N=28774)<br>Mean(SD);N or<br>Count(%) | SMD |
| **Demographics** |  |  |  |  |  |  |  |  |
| Male gender | 42199(55.79%) | 17136(59.55%) | 25063(53.48%) | 0.12 | 33876(58.86%) | 17136(59.55%) | 16740(58.17%) | 0.03 |
| Female gender | 33439(44.20%) | 11638(40.44%) | 21801(46.51%) | 0.12 | 23672(41.13%) | 11638(40.44%) | 12034(41.82%) | 0.03 |
| Baseline age, years | 62.3(12.8);n=75638 | 57.9(11.3);n=28774 | 65.1(12.9);n=46864 | 0.59* | 58.5(11.3);n=57548 | 57.9(11.3);n=28774 | 59.2(11.1);n=28774 | 0.12 * |
| Financial aids | 2750(3.63%) | 1167(4.05%) | 1583(3.37%) | 0.04 | 2070(3.59%) | 1167(4.05%) | 903(3.13%) | 0.05 |
| **Past comorbidities** |  |  |  |  |  |  |  |  |
| Charlson standard comorbidity index | 2.0(1.5);n=75638 | 1.6(1.3);n=28774 | 2.3(1.6);n=46864 | 0.52* | 1.6(1.3);n=57548 | 1.6(1.3);n=28774 | 1.7(1.3);n=28774 | 0.06 |
| Duration from earliest diabetes mellitus diagnosis date to baseline date, day | 6.5(4.9);n=75638 | 6.49(4.87);n=28774 | 6.5(4.98);n=46864 | <0.01 | 6.5(4.9);n=57548 | 6.5(4.9);n=28774 | 6.4(4.9);n=28774 | 0.01 |
| Number of hospitalizations | 1.2(0.8);n=75638 | 1.3(1.1);n=28774 | 1.2(0.6);n=46864 | 0.17 | 1.3(1.0);n=57548 | 1.3(1.1);n=28774 | 1.2(0.9);n=28774 | 0.1 |
| Hyperlipidaemia | 55015(72.73%) | 21874(76.02%) | 33141(70.71%) | 0.12 | 43882(76.25%) | 21874(76.02%) | 22008(76.48%) | 0.01 |
| Hypertension | 40638(53.72%) | 16910(58.76%) | 23728(50.63%) | 0.16 | 33575(58.34%) | 16910(58.76%) | 16665(57.91%) | 0.03 |
| Cardiac dysrhythmia | 3203(4.23%) | 984(3.41%) | 2219(4.73%) | 0.07 | 1945(3.37%) | 984(3.41%) | 961(3.33%) | <0.01 |
| VT/VF/SCD | 187(0.24%) | 92(0.31%) | 95(0.20%) | 0.02 | 180(0.31%) | 92(0.31%) | 88(0.30%) | <0.01 |
| Stroke/transient<br>ischemic attack | 2397(3.16%) | 740(2.57%) | 1657(3.53%) | 0.06 | 1456(2.53%) | 740(2.57%) | 716(2.48%) | 0.01 |
| Heart failure | 2458(3.24%) | 748(2.59%) | 1710(3.64%) | 0.06 | 1488(2.58%) | 748(2.59%) | 740(2.57%) | <0.01 |
| Ischemic heart<br>disease | 7777(10.28%) | 3616(12.56%) | 4161(8.87%) | 0.12 | 6909(12.00%) | 3616(12.56%) | 3293(11.44%) | 0.03 |
| Acute myocardial<br>infarction | 2142(2.83%) | 955(3.31%) | 1187(2.53%) | 0.05 | 1885(3.27%) | 955(3.31%) | 930(3.23%) | <0.01 |
| Peripheral<br>vascular disease | 540(0.71%) | 151(0.52%) | 389(0.83%) | 0.04 | 301(0.52%) | 151(0.52%) | 150(0.52%) | <0.01 |
| Acquired valve<br>disease | 408(0.53%) | 167(0.58%) | 241(0.51%) | 0.01 | 332(0.57%) | 167(0.58%) | 165(0.57%) | <0.01 |
| Chronic rheumatic<br>heart disease | 195(0.25%) | 85(0.29%) | 110(0.23%) | 0.01 | 170(0.29%) | 85(0.29%) | 85(0.29%) | <0.01 |
| Hypertrophic<br>cardiomyopathy | 62(0.08%) | 26(0.09%) | 36(0.07%) | <0.01 | 52(0.09%) | 26(0.09%) | 26(0.09%) | <0.01 |
| Peripheral arterial<br>disease | 478(0.63%) | 140(0.48%) | 338(0.72%) | 0.03 | 277(0.48%) | 140(0.48%) | 137(0.47%) | <0.01 |
| Systemic<br>embolism | 118(0.15%) | 39(0.13%) | 79(0.16%) | 0.01 | 78(0.13%) | 39(0.13%) | 39(0.13%) | <0.01 |
| Prosthetic valve<br>replacement or<br>valve repair | 22(0.02%) | 10(0.03%) | 12(0.02%) | 0.01 | 20(0.03%) | 10(0.03%) | 10(0.03%) | <0.01 |
| Haematological<br>malignancy | 56(0.07%) | 22(0.07%) | 34(0.07%) | <0.01 | 44(0.07%) | 22(0.07%) | 22(0.07%) | <0.01 |
| Cancer | 1995(2.63%) | 585(2.03%) | 1410(3.00%) | 0.06 | 1156(2.00%) | 585(2.03%) | 571(1.98%) | <0.01 |
| Liver diseases | 3049(4.03%) | 1369(4.75%) | 1680(3.58%) | 0.06 | 2650(4.60%) | 1369(4.75%) | 1281(4.45%) | 0.01 |
| Autoimmune<br>disease | 797(1.05%) | 312(1.08%) | 485(1.03%) | <0.01 | 618(1.07%) | 312(1.08%) | 306(1.06%) | <0.01 |
| Gastrointestinal<br>bleeding | 1735(2.29%) | 524(1.82%) | 1211(2.58%) | 0.05 | 1036(1.80%) | 524(1.82%) | 512(1.77%) | <0.01 |
| Renal diseases | 6054(8.00%) | 1535(5.33%) | 4519(9.64%) | 0.16 | 3010(5.23%) | 1535(5.33%) | 1475(5.12%) | 0.01 |
| Immunodeficiency | 11(0.01%) | 3(0.01%) | 8(0.01%) | 0.01 | 6(0.01%) | 3(0.01%) | 3(0.01%) | <0.01 |
| Diabetic retinopathy | 5277(6.97%) | 1951(6.78%) | 3326(7.09%) | 0.01 | 3776(6.56%) | 1951(6.78%) | 1825(6.34%) | 0.02 |
| Diabetic nephropathy | 922(1.21%) | 172(0.59%) | 750(1.60%) | 0.1 | 342(0.59%) | 172(0.59%) | 170(0.59%) | <0.01 |
| Diabetic neuropathy | 1830(2.41%) | 677(2.35%) | 1153(2.46%) | 0.01 | 1340(2.32%) | 677(2.35%) | 663(2.30%) | <0.01 |
| **Medications** |  |  |  |  |  |  |  |  |
| Number of anti-diabetic drug | 4.7(1.8);n=75638 | 4.8(1.5);n=28774 | 4.6(1.9);n=46864 | 0.08 | 4.7(34.0);n=57548 | 4.7(35.0);n=28774 | 4.7(32.9);n=28774 | <0.01 |
| Metformin | 67930(89.80%) | 26864(93.36%) | 41066(87.62%) | 0.2 | 53807(93.49%) | 26864(93.36%) | 26943(93.63%) | 0.01 |
| Sulphonylurea | 58107(76.82%) | 20689(71.90%) | 37418(79.84%) | 0.19 | 42614(74.04%) | 20689(71.90%) | 21925(76.19%) | 0.1 |
| Insulin | 38542(50.95%) | 14795(51.41%) | 23747(50.67%) | 0.01 | 29644(51.51%) | 14795(51.41%) | 14849(51.60%) | <0.01 |
| Acarbose | 2563(3.38%) | 1032(3.58%) | 1531(3.26%) | 0.02 | 1995(3.46%) | 1032(3.58%) | 963(3.34%) | 0.01 |
| Thiozolidinedone | 15186(20.07%) | 7852(27.28%) | 7334(15.64%) | 0.29* | 14518(25.22%) | 7852(27.28%) | 6666(23.16%) | 0.1 |
| Glucagon-like peptide-1 receptor agonists | 3963(5.23%) | 1413(4.91%) | 2550(5.44%) | 0.02 | 2804(4.87%) | 1413(4.91%) | 1391(4.83%) | <0.01 |
| ACEI/ARB | 21841(28.87%) | 9143(31.77%) | 12698(27.09%) | 0.1 | 18972(32.96%) | 9143(31.77%) | 9829(34.15%) | 0.05 |
| Anticoagulants | 37474(49.54%) | 16665(57.91%) | 20809(44.40%) | 0.27* | 34102(59.25%) | 16665(57.91%) | 17437(60.59%) | 0.05 |
| Antiplatelets | 14435(19.08%) | 6162(21.41%) | 8273(17.65%) | 0.09 | 11455(19.90%) | 6162(21.41%) | 5293(18.39%) | 0.08 |
| Statins and fibrates | 39011(51.57%) | 16581(57.62%) | 22430(47.86%) | 0.2 | 32493(56.46%) | 16581(57.62%) | 15912(55.29%) | 0.05 |
| Other lipid-lowering drugs | 25887(34.22%) | 10129(35.20%) | 15758(33.62%) | 0.03 | 22061(38.33%) | 10129(35.20%) | 11932(41.46%) | 0.13 * |
| Systemic steroids | 611(0.80%) | 241(0.83%) | 370(0.78%) | 0.01 | 482(0.83%) | 241(0.83%) | 241(0.83%) | <0.01 |
| Diuretics | 33372(44.12%) | 16236(56.42%) | 17136(36.56%) | 0.41* | 30539(53.06%) | 16236(56.42%) | 14303(49.70%) | 0.13 |
| Alpha-blockers | 9439(12.47%) | 3158(10.97%) | 6281(13.40%) | 0.07 | 6006(10.43%) | 3158(10.97%) | 2848(9.89%) | 0.04 |
| Beta-blockers | 14178(18.74%) | 7041(24.47%) | 7137(15.22%) | 0.23* | 13049(22.67%) | 7041(24.47%) | 6008(20.87%) | 0.09 |
| Calcium channel blockers (dihydropyridine) | 10298(13.61%) | 5304(18.43%) | 4994(10.65%) | 0.22* | 10160(17.65%) | 5304(18.43%) | 4856(16.87%) | 0.04 |
| Calcium channel blocker (non-dihydropyridine) | 477(0.63%) | 218(0.75%) | 259(0.55%) | 0.03 | 434(0.75%) | 218(0.75%) | 216(0.75%) | <0.01 |
| Non-steroidal anti-inflammatory drugs | 13528(17.88%) | 5091(17.69%) | 8437(18.00%) | 0.01 | 10275(17.85%) | 5091(17.69%) | 5184(18.01%) | 0.01 |
| **Calculated biomarkers** |  |  |  |  |  |  |  |  |
| Abbreviated MDRD, mL/min/1.73m<sup>2</sup> | 80.8(28.4);n=62788 | 89.8(24.1);n=24207 | 75.2(29.4);n=38581 | 0.54* | 87.0(25.5);n=48387 | 89.8(24.1);n=24207 | 88.2(26.5);n=24180 | 0.12 * |
| Prognostic nutritional index | 41.1(6.3);n=49625 | 41.9(5.9);n=20758 | 40.5(6.5);n=28867 | 0.22* | 41.7(6.0);n=40574 | 41.9(5.9);n=20758 | 41.5(6.1);n=19816 | 0.07 |
| **Complete blood counts** |  |  |  |  |  |  |  |  |
| White cell count, x10<sup>9</sup>/L | 8.0(2.9);n=39444 | 7.99(2.64);n=16396 | 8.04(3.14);n=23048 | 0.02 | 8.0(3.1);n=32206 | 7.99(2.64);n=16396 | 8.03(3.45);n=15810 | 0.02 |
| **Lipid profiles** |  |  |  |  |  |  |  |  |
| Triglyceride, mmol/L | 1.7(1.6);n=58972 | 1.8(1.8);n=23142 | 1.7(1.4);n=35830 | 0.08 | 1.8(1.7);n=46235 | 1.82(1.8);n=23142 | 1.83(1.66);n=23093 | <0.01 |
| Low-density lipoprotein, mmol/L | 2.4(0.8);n=57905 | 2.39(0.81);n=22704 | 2.39(0.8);n=35201 | 0.01 | 2.4(0.8);n=45234 | 2.39(0.81);n=22704 | 2.41(0.82);n=22530 | 0.03 |
| High-density lipoprotein, mmol/L | 1.2(0.3);n=58879 | 1.17(0.31);n=23101 | 1.21(0.34);n=35778 | 0.14 | 1.2(0.3);n=46154 | 1.17(0.31);n=23101 | 1.18(0.33);n=23053 | 0.04 |
| Total cholesterol, mmol/L | 4.3(1.0);n=59033 | 4.35(1.02);n=23171 | 4.34(0.99);n=35862 | 0.01 | 4.4(1.0);n=46280 | 4.3(1.0);n=23171 | 4.4(1.0);n=23109 | 0.04 |
| mmol/L |  |  |  |  |  |  |  |  |
| **Glucose tests and variabilities** |  |  |  |  |  |  |  |  |
| Haemoglobin A1C, % | 8.1(1.6);n=61542 | 8.3(1.6);n=23793 | 8.0(1.5);n=37749 | 0.21* | 8.3(1.6);n=47544 | 8.3(1.6);n=23793 | 8.2(1.7);n=23751 | 0.04 |
| Fasting glucose, mmol/L.1 | 9.0(3.9);n=55823 | 9.2(3.7);n=21892 | 8.8(4.0);n=33931 | 0.1 | 9.2(3.8);n=43855 | 9.22(3.68);n=21892 | 9.21(3.96);n=21963 | <0.01 |

### Primary analysis

In the matched cohort, 104 SGLT2I users and 161 DPP4I users developed infective endocarditis. After a follow-up median of 5.6 years, the incidence of infective endocarditis was lower amongst SGLT2I users (Incidence: 0.65 per 1000 person-year; 95% Confidence interval [CI]: 0.53-0.79] than in DPP4I users (Incidence: 1.04 per 1000 person-year; 95% CI: 0.89-1.21). SGLT2I was associated with a 42% (Hazard ratio [HR]: 0.58; 95% CI: 0.41-0.81) lower risks of infective carditis compared to DPP4I after adjustments **(Table 2; Supplementary Table 2)**. This was substantiated by the cumulative incidence curves stratified by SGLT2I versus DPP4I **(Figure 2)**. The marginal effects plots demonstrated that SGLT2I was associated with lower risks of infective endocarditis regardless of the duration of the diabetes diagnosis prior drug uses and the MDRD (**Supplementary Figure 3**).

**Figure 2.**
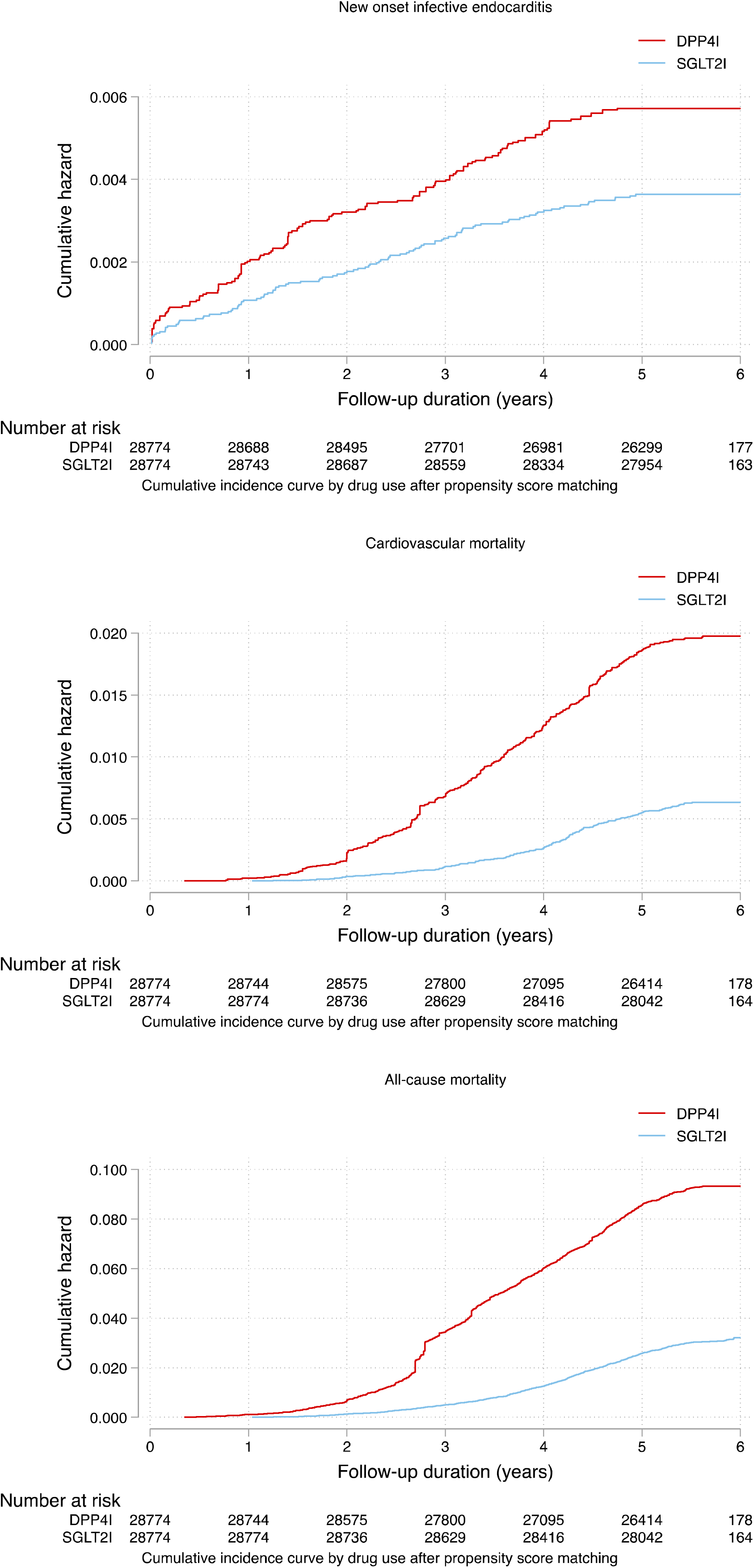
Cumulative incidence curves for new onset infective endocarditis, cardiovascular mortality, and all-cause mortality stratified by drug exposure effects of SGLT2I and DPP4I after propensity score matching (1:1). SGLT2I: Sodium-glucose cotransporter-2 inhibitors; DPP4I: Dipeptidyl peptidase-4 inhibitors.

**Table 2.** Incidence rate (IR) per 1000 person-year and adjusted hazard ratio of new onset infective endocarditis, cardiovascular mortality, and all-cause mortality in the cohort before and after 1:1 propensity score matching.

| **Infective endocarditis** | **Number of patients** | **Number of events** | **Total person-year** | **Follow-up duration, years [95% CI]** | **Incidence [95% CI] per 1000 person-year** | **Adjusted hazard ratio [95% CI]** | **P-value** |
| --- | --- | --- | --- | --- | --- | --- | --- |
| **DPP4I** | 28774 | 161 | 154689.3 | 5.57 [5.26-5.8] | 1.04 [0.89-1.21] | 1 [reference] | NA |
| **SGLT2I** | 28774 | 104 | 159279.1 | 5.62 [5.35-5.83] | 0.65 [0.53-0.79] | 0.58 [0.41-0.81] | 0.0016 |
| **Cardiovascular-related mortality** | **Number of patients** | **Number of events** | **Total person-year** | **Follow-up duration, years [95% CI]** | **Incidence [95% CI]** | **Adjusted hazard ratio [95% CI]** | **P value** |
| **DPP4I** | 28774 | 538 | 155170 | 5.57 [5.27-5.8] | 3.47 [3.18-3.77] | 1 [reference] | NA |
| **SGLT2I** | 28774 | 176 | 159614.9 | 5.62 [5.35-5.83] | 1.10 [0.95-1.27] | 0.49[0.33-0.72] | 0.0002 |
| **All-cause mortality** | **Number of patients** | **Number of events** | **Total person-year** | **Follow-up duration, years [95% CI]** | **Incidence [95% CI]** | **Adjusted hazard ratio [95% CI]** | **P value** |
| **DPP4I** | 28774 | 2524 | 155170 | 5.57 [5.27-5.8] | 16.3 [15.6-16.9] | 1 [reference] | NA |
| **SGLT2I** | 28774 | 851 | 159614.9 | 5.62 [5.35-5.83] | 5.33 [4.98-5.70] | 0.42 [0.29-0.62] | <0.0001 |

**Table 3.**
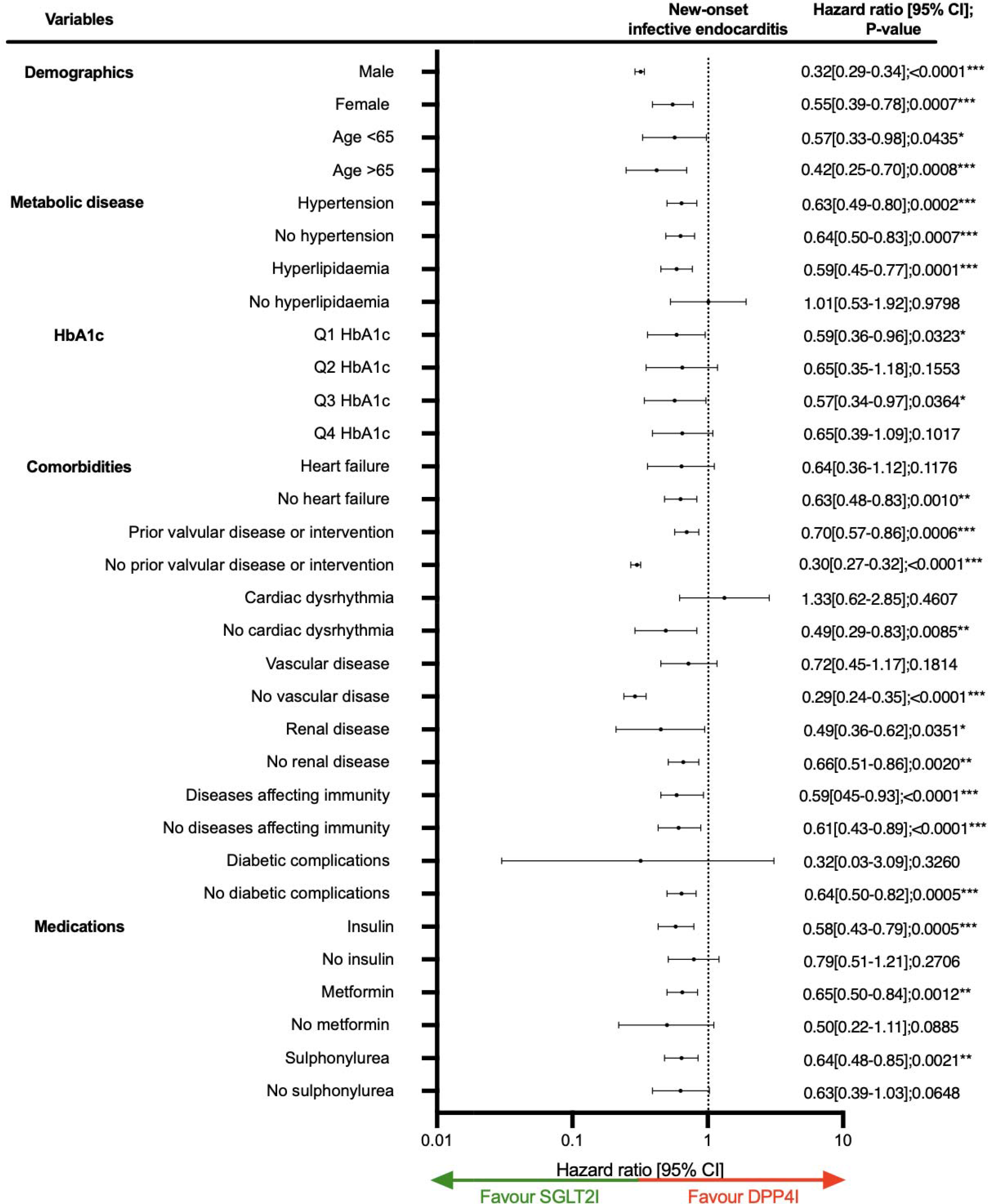
Forests plot of hazard ratios with 95% CI of subgroup analysis of SGLT2I v.s. DPP4I effects on new onset infective endocarditis in patients with type-2 diabetes mellitus. * for p≤ 0.05, ** for p ≤ 0.01, *** for p ≤ 0.001; HR: hazard ratio; CI: confidence interval; SGLT2I: sodium glucose cotransporter-2 inhibitor; DPP4I: dipeptidyl peptidase-4 inhibitor; MDRD: modification of diet in renal disease; Vascular disease: patients with stroke/transient ischemic attack, ischemic heart disease, acute myocardial infarction, and peripheral vascular disease Diabetic complications: patients with diabetic retinopathy, neuropathy and nephropathy Diseases affecting immunity: patients on systemic steroids, with immunodeficiency, haematological malignancies, cancer, and autoimmune diseases Prior valvular disease or intervention: Patient with acquired valve disease, chronic rheumatic heart disease, prosthetic valve replacement or valve repair

After a follow-up of 5.6 years, 538 DPP4I and 176SGLT2I users passed away due to cardiovascular diseases. The incidence of cancer-related mortality was lower amongst SGLT2I users (Incidence: 1.10 per 1000 person-year; 95% CI:0.95-1.27) compared to DPP4I users (Incidence: 3.47 per 1000 person-year; 95% CI: 3.18-3.77). SGLT2I was associated with a 51% lower risks of cardiovascular diseases related mortality compared to DPP4I. Furthermore, 851 SGLT2I users and 2524 DPP4I users passed away. The incidence of all-cause mortality was lower amongst SGLT2I users (Incidence: 5.33 per 1000 person-year; 95% CI: 4.98-5.70) compared to DPP4I users (Incidence: 16.3 per 1000 person-year; 95% CI: 15.6-16.9). SGLT2I was associated with a 58% lower risks of all-cause mortality compared to DPP4I users. The marginal effects plot demonstrated the effects regardless of the duration of the diabetes diagnosis prior drug uses and the MDRD (**Supplementary Figure 3**).

### Subgroup analysis

The results of the subgroup analysis for the effects of SGLT2I and DPP4I on infective endocarditis are shown in **Figure 3**. In the subgroup analysis, the results demonstrated that SGLT2I was associated with lower risks of infective endocarditis regardless of age and gender. The results were consistent amongst patients with the first and third quartile of HbA1c, while unsignificant in the remaining 2 quartiles because of insufficient power in the subgroup. The association remained significant regardless of hypertension, prior valvular disease or intervention, renal diseases, diseases affecting immunity. Meanwhile, the association were only significant amongst patients with hyperlipidaemia, without heart failure, without cardiac arrhythmia, without vascular disease, and without diabetic complications. Furthermore, the association were significant only amongst patient on insulin, metformin, or sulphonylurea; those could be due to the insufficient sample size in the subgroups amongst the patients with the diseases or drug users.

### Sensitivity analysis

Sensitivity analyses were performed to confirm the predictability of the models. The results of the cause-specific hazard models, sub-distribution hazard models and different propensity score approaches demonstrated that different models do not change the point estimates for both the primary and the secondary outcomes (all P<0.05) **(Supplementary Table 3).** A 3-arm analysis with the inclusion of GLP1a including patients only on SGLT2I, DPP4I, and GLP1a using stabilized IPTW was conducted (**Supplementary Table 4**). The results between DPP4I and SGLT2I remained consistent with the main result (p<0.05) **(Table 2)**. However, it was found that SGLT2I was not associated with lower risks of prostate cancer compared to GLP1a (HR: 0.81; 95% CI: 0.62-1.07). Regarding patients with potential contraindication due to renal diseases, upon excluding patients with CKD stage 4/5 (eGFR <30 mL/min/1.73m^2), peritoneal dialysis or haemodialysis in the matched cohort, it was demonstrated that SGLT2I was associated with lower risks of infective endocarditis compared to DPP4I **(Supplementary Table 5)**. Besides, the association remained significant after excluding patients with drug abuse, excluding patients with financial aid, and including patients developed infective endocarditis within 30 days after medications initiation (p<0.05) **(Supplementary Table 6)**. The sensitivity analysis for 1-year lag time also demonstrated the same trend (p<0.05) **(Supplementary Table 7)**. Lastly, to confirm the association with the laboratory tests, when defining new-onset infective endocarditis using only two separate positive blood culture for infective endocarditis microorganisms, 41 (Incidence: 26.43 per 100000 person-year; 95% CI: 18.98-35.87) and 22 DPP4I users (Incidence: 14.19 per 100000 person-year; 95% CI: 9.92-21.48) developed infective endocarditis during follow-up; SGLT2I remained to be associated with lower risks of infective endocarditis (HR: 0.57; 95% CI: 0.34-0.95) compared to DPP4I after adjustments **(Supplementary Table 8)**.

## Discussion

In this territory-wide cohort study, we utilised real-world data to compare the association between SGLT2I, DPP4I and new-onset infective endocarditis. Our results suggest that SGLT2I usage is associated with a lower risk of new-onset infective endocarditis than DPP4I usage. As of now, this is the first study to examine the effects of two anti-diabetic medications and demonstrate their potential effects on preventing infective endocarditis.

### Comparison with previous studies

Patients with diabetes often suffer from an increased susceptibility to infectious diseases. As infective endocarditis remains a relatively rare condition compared to other major infections, there is limited evidence surrounding its prognosis and prevalence amongst diabetic patients. The incidence of infective endocarditis in our study was higher than the results of existing literature regarding the incidence in the general population in Hong Kong (IR: 4.9 [4.8-5.1] per 100,000 person-year).^20^ As such, a sensitivity analysis was conducted **(Supplementary Table 8)** by defining the infective using the blood culture results in order to ensure the association was not due to over-diagnosis of infective endocarditis. In this sensitivity analysis, the association remained consistent with similar adjusted hazard ratio and the incidence remained higher than the described literature. In Hong Kong diabetic population, *Streptococci* and *Staphylococcus aureus* was the most common pathogen in patients with infective endocarditis.^24^ It was previously suggested that T2DM was associated with increased risks of infective endocarditis. An observational study conducted in Spain reported an increased in infective endocarditis incidence amongst diabetic patients compared to non-diabetic patients.^3^ Furthermore, the incidence of IE has been increasing across the years.^3, 25^ It was suggested that amongst T2DM patients with a duration of DM 5-10 years the incidence can be up to 0.33 per 1000 person-year regardless of extent of diabetic control.^26^ This cohort included T2DM patients requiring second-line anti-diabetic drugs, signifying that the diabetic controls were relatively poor which may result in even high incidence of infective endocarditis.

Due to the high mortality rate associated with infective endocarditis,^27, 28^ seeking therapeutic strategies that may minimise the risk of infective endocarditis becomes imperative to optimise management of patients with diabetes. Some studies have demonstrated that SGLT2I exhibit pleiotropic effects against pneumonia (HR: 0.63; 95% CI: 0.55-0.72), sepsis (HR: 0.52; 95% CI: 0.44-0.62), and other infectious diseases.^29, 30^ A meta-analysis suggests that SGLT2I correlated with reduced gastroenteritis compared to placebo^31^. This was further supported by Yin *et al.* showing reduced risks of bronchitis, pneumonia, respiratory tract infection amongst SGLT2I users compared to placebo.^32^ Similarly, there have been some preliminary evidence suggesting that DPP4I may be beneficial against infections.^33^ Hence, it can be hypothesized that SGLT2I and DPP4I may exert potential protective effects against infective endocarditis in patients with diabetes. However, this current association is not well-defined as there is little to no evidence corroborating this claim.

To the best of our knowledge, there is currently no existing clinical trial study that compares the effects of SGLT2I and DPP4I on infective endocarditis. Interestingly, some studies show contrasting results, suggesting that the use of SGLT2I and DPP4I may increase risk of infection.^34, 35^ Further prospective investigation is crucial to comprehend the mechanism associated with SGLT2I and infective endocarditis. There are currently no definitive claims that can explain the complex mechanism behind the pleotropic effects of SGLT2I on infective endocarditis. One explanation postulated that anti-diabetic agents are related to oxidative stress, which may protect against the production of hydrogen peroxide and thiobarbituric acid-reactive substances in patients with active infective endocarditis.^36, 37^

### Clinical implications

In recent years, the potential protective effects of SGLT2I and DPP4I on various infectious diseases have received worldwide attention. The study data was obtained from routine clinical practice in Hong Kong and suggests that SGLT2I may show more significant protective effects against infective endocarditis compared to DPP4I amongst T2DM patients. As the mortality and incidence trends of infective endocarditis have continued to escalate globally,^38^ the findings of this study could provide vital information for refining treatment of diabetes and taking precautions to avoid adverse drug effects. Therefore, it is prudent to include more randomised clinical trials and observational studies that scrutinize the inherent mechanism and drug-drug interactions of SGLT2I and DPP4I usage. This notwithstanding, this may also amend current clinical guidelines that govern treatment of infective endocarditis and diabetes, allowing clinicians to make more informed decisions when recommending different types of diabetes treatment.

### Limitations

In this study, several limitations should be acknowledged. Firstly, due to its observational nature, information on predictive variables such as smoking and dental hygiene, as well as diagnostic data such as echocardiogram, were not available from CDARS.^39^ To address this, extensive laboratory results and comorbidities related to infective endocarditis, such as illicit drug uses, financial status, cardiac procedures, valvular heart diseases, and immunocompromised, were included to infer for possible risk variables. Moreover, our cohorts had been well matched across a range of medications and diseases, adjusted the regression, and performed a variety of sensitivity analyses to minimise the impact of the bias. Secondly, the observational nature rendered the data susceptible to under-coding, missing data and coding errors which could result in information bias. The patient’s drug exposure duration and level of compliance could not be standardised. Furthermore, due to the relatively small sample size of the GLP1a cohort, the association between SGLT2I and GLP1a could be insignificant due to insufficient power; further studies are needed as GLP1a continued to gain popularity in Hong Kong.^40^ Lastly, the results from this study are correlational in nature, and further studies should be conducted to examine the anti-infective effects of SGLT2I.

## Conclusion

In this population-based cohort study, SGLT2I use was associated with lower risks of new-onset infective endocarditis compared to DPP4I amongst T2DM patients after adjustments regardless of prior valvular diseases, cardiovascular diseases, and immune status. Further randomised controlled trials and mechanistic studies are needed to confirm the association.

## Data Availability

An anonymised version without identifiable or personal information is available from the corresponding authors upon reasonable request for research purposes.

## Funding source

The authors received no funding for the research, authorship, and/or publication of this article.

## Ethical approval statement

This study was approved by the Institutional Review Board of the University of Hong Kong/Hospital Authority Hong Kong West Cluster (HKU/HA HKWC IRB) (UW-20-250) and complied with the Declaration of Helsinki.

## Conflicts of Interest

None.

## Acknowledgements

None.

## Guarantor Statement

All authors approved the final version of the manuscript. GT is the guarantor of this work and, as such, had full access to all the data in the study and takes responsibility for the integrity of the data and the accuracy of the data analysis.

## Author contributions

Data analysis: OHIC, TG, FJ, JSKC, JZ

Data review: OHIC, FJ, DIS, SL, GT, JZ

Data acquisition: OHIC, SL, GT

Data interpretation: OHIC, CTC, BMYC, GT, JZ

Critical revision of manuscription: FJ, RTKP, WTW, GYHL, BMYC, GT, JZ

Supervision: GYHL, BMYC, GT, JZ

Manuscript writing: OHIC, CTC, JSKC, DIS

Manuscript revision: OHIC, CTC, JSKC, DIS, RTKP

## Notes

### Competing Interest Statement

The authors have declared no competing interest.

