## Supplementary Appendix for "New-onset infective endocarditis in diabetic patients receiving SGLT2I, DPP4I and GLP1a: A population-based cohort study"

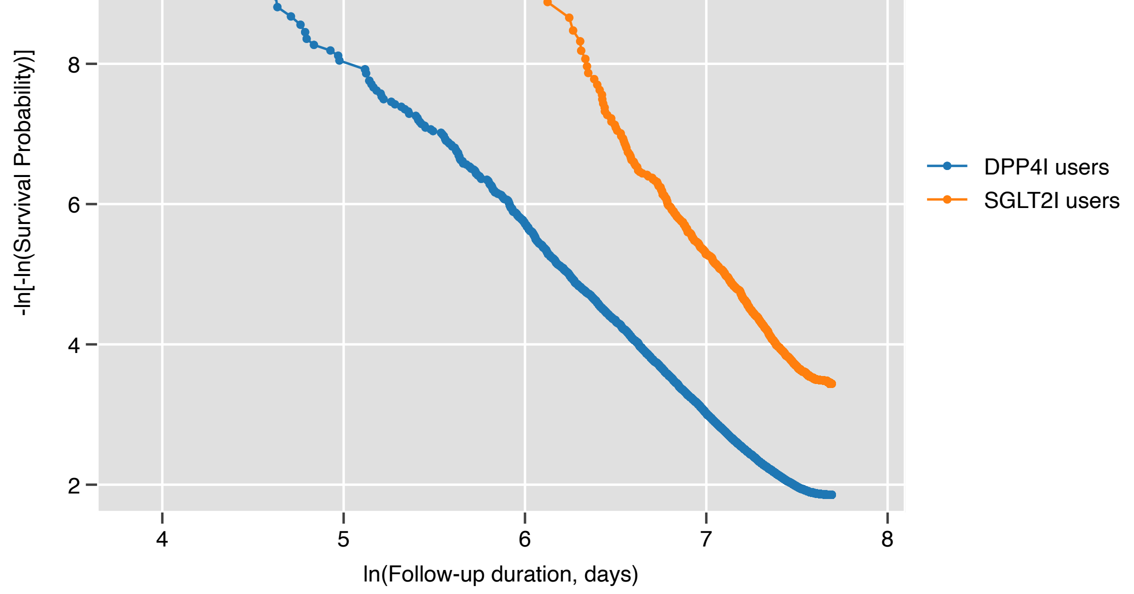

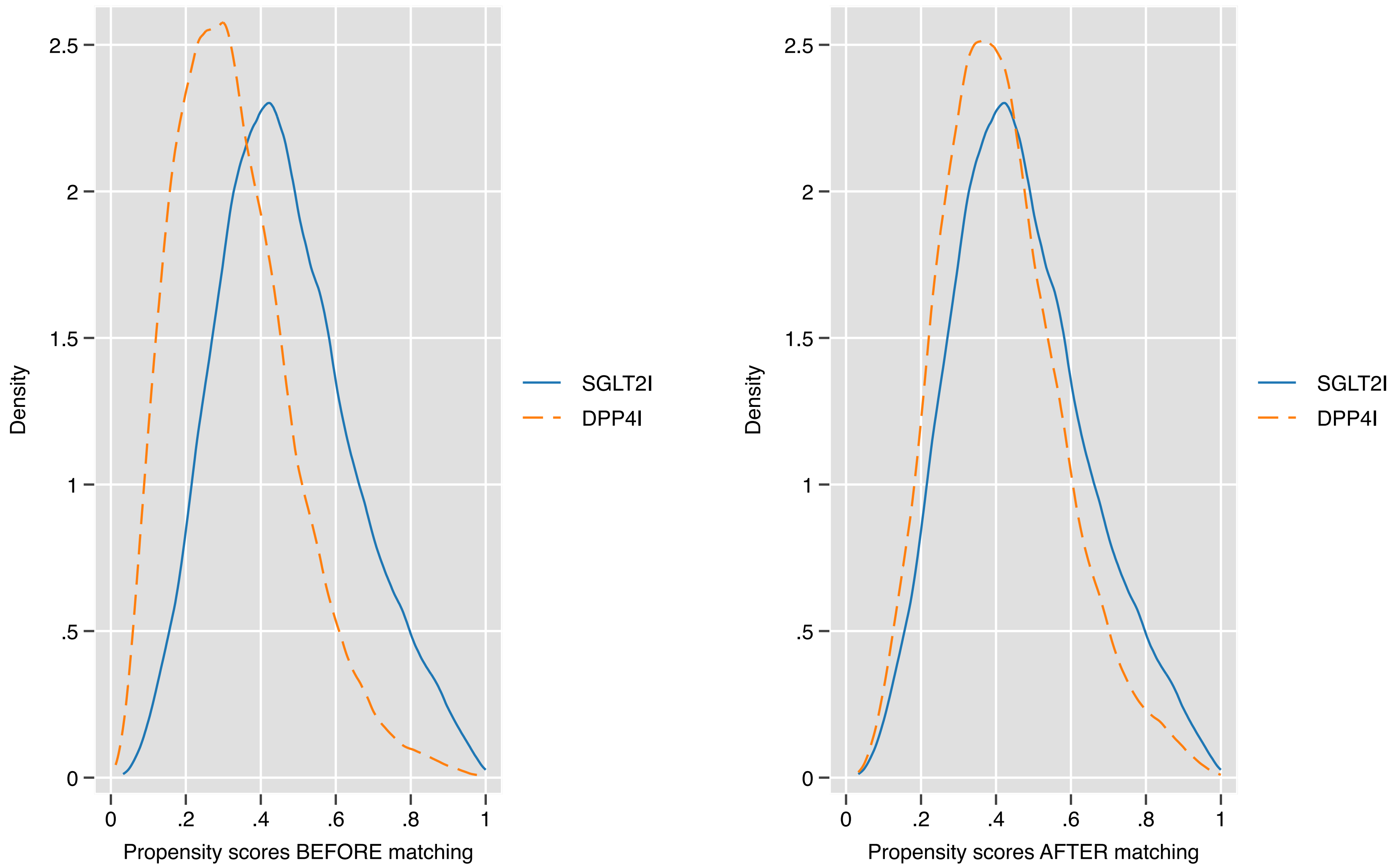

Nearest neighbor search strategy with caliper=0.1.

**Supplementary Figure 1. Propensity score matching comparisons and proportional hazard assumption checking with parallel lines for SGLT2I v.s. DPP4I before and after 1:1 matching with nearest neighbour search strategy with calliper of 0.1**

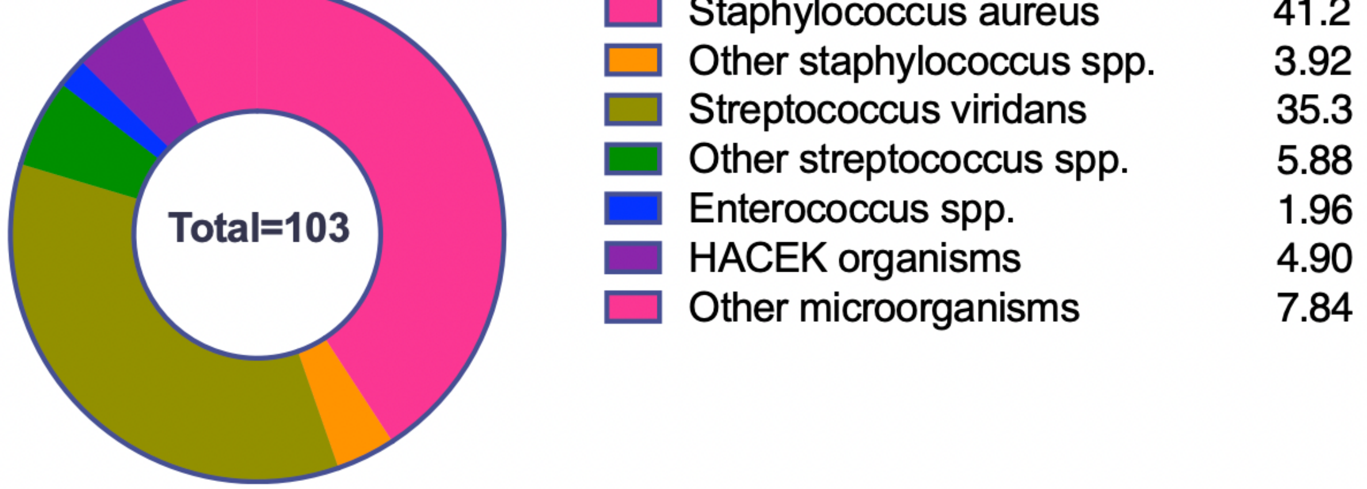

**Supplementary Figure 2. The pathogens responsible for the new-onset infective endocarditis amongst the patients with identified blood culture results after propensity score matching (1:1).**

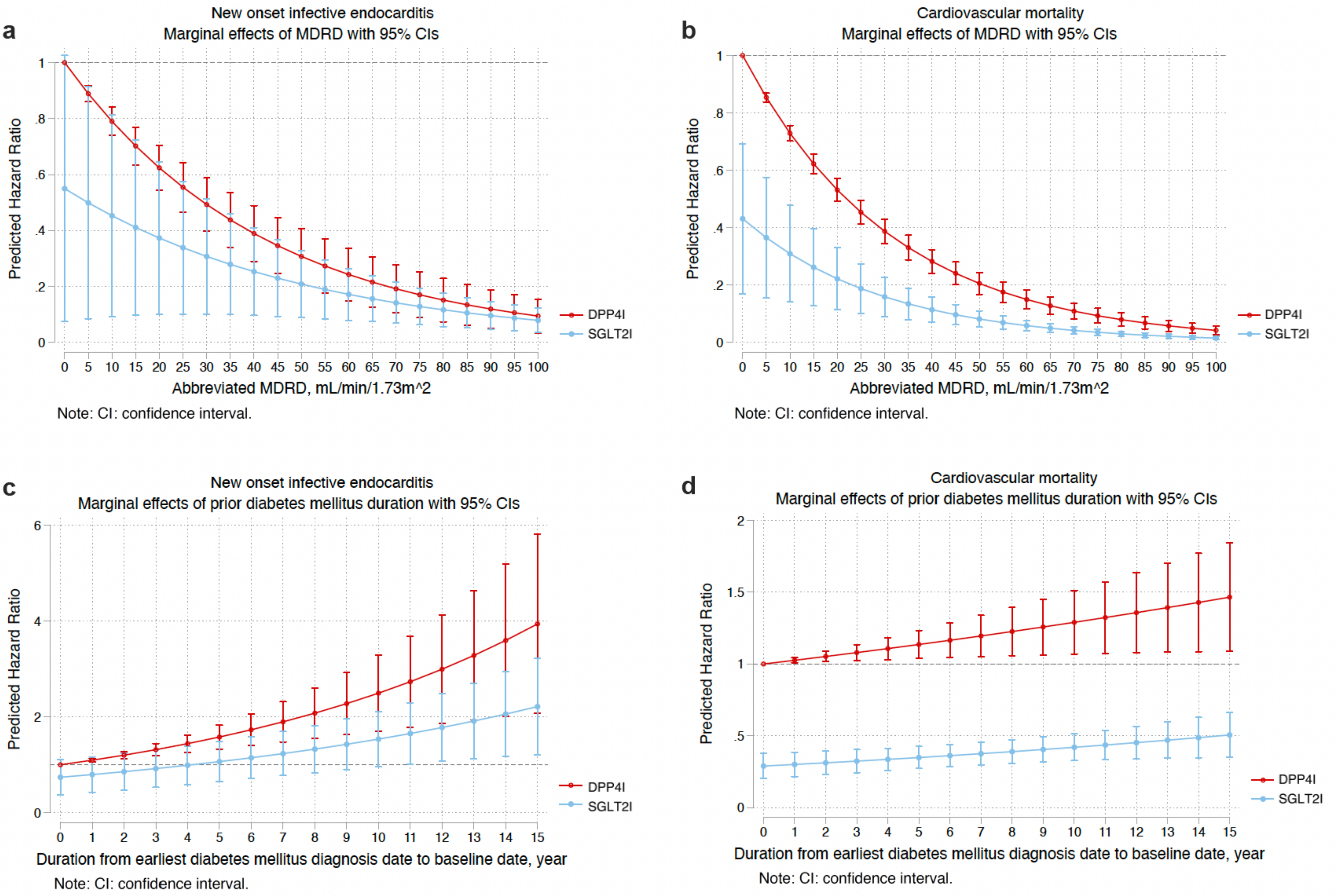

**ry Figure 3. Marginal effects of MARD, number of prior anti-diabetic drugs, and prior diabetes duration with 95% Cis on new onset infective endocarditis, cardiovascular mo tality stratified by drug use in the matched cohort.**

m-glucose cotransporter-2 inhibitors; DPP4I: Dipeptidyl peptidase-4 inhibitors.

**Supplementary Table 1. The International Classification of Diseases, Clinical Modification (ICD-9-CM) codes for definitions of past comorbidities and outcomes.**

| Adverse outcome of interest |
|---|
| **Infective endocarditis:** 036.42 098.84 112.81 115.04 115.14 115.94 421 424.9x OR 2 separate positive blood culture for typical infective endocarditis organisms |
| **Past comorbidities** |
| **Cancer**: 140-239 |
| **Hypertension**: 401 401.1 401.9 402 402.01 402.1 402.11 402.9 402.91 403 403.01 403.1 403.11 403.9 403.91 404 404.01 404.02 404.03 404.1 404.11 404.12 404.13 404.9 404.91 404.92 404.93 405 405.01 405.09 405.1 405.11 405.19 405.9 405.91 405.99 437.2 |
| **Hyperlipidaemia**: 272.0 272.1 272.2 272.3 272.4 |
| **Liver diseases**: 571.0 571.1 571.2 571.3 571.40 571.41 571.42 471.49 571.5 571.6 571.8, 571.9 |
| **Autoimmune diseases:** 136.1, 359.79, 359.71, 443.1, 446, 555, 556.8~556.9, 556.0~556.6, 695.4, 710, 714, 720, 725 |
| **Gastrointestinal bleeding:** 578.9 |
| **Heart failure:** 428 428 428.1 428.2 428.2 428.21 428.22 428.23 428.3 428.3 428.31 428.32 428.33 428.4 428.4 428.41 428.42 428.43 428.9 398.91 402.01 402.11 402.91 404.01 404.03 404.11 404.13 404.91 404.93 |
| **Atrial fibrillation:** 427.31 429.4 |
| **Stroke/transient ischemic attack:** 435 435.1 435.2 435.3 435.8 435.9 433.81 433.91 434 436 437 437.1 433.31 433.01 434.01 434.1 434.11 434.9 434.91 437.2 437.3 437.4 437.5 437.6 437.7 437.8 437.9 430 431 432 432.1 432.9 |
| **Ischemic heart disease:** 410.01 410.02 410.1 410.11 410.12 410.2 410.21 410.22 410.3 410.31 410.32 410.4 410.41 410.42 410.5 410.51 410.52 410.6 410.61 410.62 410.7 410.71 410.72 410.8 410.81 410.82 410.9 410.91 410.92 411 411.1 411.8 411.81 411.89 413 413.1 413.9 414 414.01 414.02 414.03 414.04 414.05 414.06 414.07 414.1 414.11 414.12 414.19 414.2 414.3 414.4 414.8 414.9 410 412 |
| **Peripheral vascular disease:** 250.7 443.9 443 443.1 443.2 443.21 443.22 443.23 443.24 443.29 443.8 443.81 443.82 443.89 441 443.9 785.4 V43.4 |
| **Acute myocardial infarction:** 410 410.01 410.02 410.1 410.11 410.12 410.2 410.21 410.22 410.3 410.31 410.32 410.4 410.41 410.42 410.5 410.51 410.52 410.6 410.61 410.62 410.7 410.71 410.72 410.8 410.81 410.82 410.9 410.91 410.92 |
| **Renal diseases:** 582 582 582.1 582.2 582.4 582.8 582.81 582.89 582.9 583 583 583.1 583.2 583.4 583.6 583.7 585 585.1 585.2 585.3 585.4 585.5 585.6 585.9 586 588 588 588.1 588.8 588.81 588.89 588.9 + Laboratory tests eGFR <60 + Haemodialysis (39.95) Peritoneal dialysis (54.98) |
| **Acquired valve disease:** 0.93.2x 394 395 396 397 424 |
| **Chronic rheumatic heart disease**: 391 392.0 393 394.x 395.x 396.x 397.x 398.x |
| **Complications due to cardiac devise**: 996.72 |
| **Hypertrophic cardiomyopathy**: 425.1 |

| |
|---|
| **Systemic embolism:** 289.59 415.1 434.1 444.x 445.x |
| **Prosthetic valve replacement or valve repair:** 35.2x 996.02 996.71 |
| **Infection of cardiac device or implant:** 996.61 |
| **Immunodeficiency**: 279 |

**Supplementary Table 2. Multivariate Cox regression models with adjustments to predict new onset infective endocarditis, cardiovascular mortality, and all-cause mortality in the matched cohort.**

* for p≤ 0.05, ** for p ≤ 0.01, *** for p ≤ 0.001; HR: hazard ratio; CI: confidence interval; SGLT2I: sodium glucose cotransporter-2 inhibitor; DPP4I: dipeptidyl peptidase-4 inhibitor.

**Model 1** adjusted for significant demographics.

**Model 2** adjusted for significant demographics, and past comorbidities.

**Model 3** adjusted for significant demographics, past comorbidities, duration from earliest diabetes mellitus date to initial drug exposure date, and number of prior hospitalizations.

**Model 4** adjusted for significant demographics, past comorbidities, duration of diabetes mellitus, and number of prior hospitalizations, number of anti-diabetic drugs, and non-SGLT2I/DPP4I medications.

**Model 5** adjusted for significant demographics, past comorbidities, duration of diabetes mellitus, and number of prior hospitalizations, number of anti-diabetic drugs, non-SGLT2I/DPP4I medications, abbreviated MDRD.

**Model 6** adjusted for significant demographics, past comorbidities, duration of diabetes mellitus, and number of prior hospitalizations, number of anti-diabetic drugs, non-SGLT2I/DPP4I medications, abbreviated MDRD, HbA1c, fasting glucose.

| **Characteristics** | **New onset infective endocarditis HR [95% CI];P value** | **Cardiovascular mortality HR [95% CI];P value** | **All-cause mortality HR [95% CI];P value** |
|---|---|---|---|
| ***Model 1*** | 0.50[0.36-0.68];<0.0001*** | 0.29[0.23-0.36];<0.0001*** | 0.27[0.22-0.34];<0.0001*** |
| ***Model 2*** | 0.51[0.37-0.70];<0.0001*** | 0.31[0.26-0.37];<0.0001*** | 0.28[0.22-0.34];<0.0001*** |
| ***Model 3*** | 0.53[0.39-0.72];0.0001*** | 0.32[0.27-0.37];<0.0001*** | 0.29[0.24-0.35];<0.0001*** |
| ***Model 4*** | 0.54[0.35-0.83];0.0049** | 0.39[0.26-0.59];<0.0001*** | 0.33[0.26-0.41];<0.0001*** |
| ***Model 5*** | 0.56[0.41-0.76];0.0002*** | 0.40[0.27-0.57];<0.0001*** | 0.33[0.24-0.45];<0.0001*** |
| ***Model 6*** | 0.58[0.41-0.81];0.0016** | 0.49[0.33-0.72];0.0002*** | 0.42[0.29-0.62];<0.0001*** |

**Supplementary Table 3. Sensitivity analyses for exposure effects of SGLT2I v.s. DPP4I on new onset infective endocarditis, cardiovascular mortality, and all-cause mortality using different models.**

* for p≤ 0.05, ** for p ≤ 0.01, *** for p ≤ 0.001; SGLT2I: Sodium-glucose cotransporter-2 inhibitors; DPP4I: Dipeptidyl peptidase-4 inhibitors; HR: hazard ratio; CI: confidence interval; PS: propensity score; IPTW: inverse probability of treatment weighting, SIPTW: stable inverse probability of treatment weighting.

| **Model** | **New onset infective endocarditis HR [95% CI];P value** | **Cardiovascular mortality HR [95% CI];P value** | **All-cause mortality HR [95% CI];P value** |
|---|---|---|---|
| Cause-specific hazard models | 0.53[0.38-0.74];0.0001*** | 0.32[0.27-0.37];<0.0001*** | 0.24[0.18-0.33];<0.0001*** |
| Sub-distribution hazard models | 0.58[0.45-0.75];<0.0001*** | 0.34[0.28-0.42];<0.0001*** | 0.28[0.19-0.40];<0.0001*** |
| PS stratification | 0.64[0.50-0.81];0.0003*** | 0.31[0.23-0.43];<0.0001*** | 0.30[0.17-0.53];<0.0001*** |
| PS with IPTW | 0.63[0.49-0.81];0.0003*** | 0.41[0.21-0.78];0.0067** | 0.32[0.27-0.38];<0.0001*** |
| PS with SIPTW | 0.66[0.51-0.85];0.0014** | 0.40[0.28-0.58];<0.0001*** | 0.37[0.28-0.48];<0.0001*** |

**Supplementary Table 4. Sensitivity analysis: Three-arm analysis results using stabilized IPTW.**

* for p≤ 0.05, ** for p ≤ 0.01, *** for p ≤ 0.001; HR: hazard ratio; CI: confidence interval; SGLT2I: sodium glucose cotransporter-2 inhibitor; DPP4I: dipeptidyl peptidase-4 inhibitor; IPTW: Inverse probability of treatment weighting

| **Drug treatment** | **New onset infective endocarditis HR[95% CI];P value** | **Cardiovascular mortality HR[95% CI];P value** | **All-cause mortality HR[95% CI];P value** |
|---|---|---|---|
| SGLT2I v.s. DPP4I | 0.63[0.48-0.83];0.0010** | 0.43[0.38-0.47];<0.0001*** | 0.36[0.32-0.40];<0.0001*** |
| SGLT2I v.s. GLP1 | 0.81[0.62-1.07];0.1397 | 0.94[0.60-1.46];0.7720 | 0.89[0.48-1.66];0.7219 |

**Supplementary Table 5. Sensitivity analysis: Excluding patients with CKD stage 4/5 (eGFR <30), peritoneal dialysis or haemodialysis in the SGLT2I v.s. DPP4I matched cohort.**

* for p≤ 0.05, ** for p ≤ 0.01, *** for p ≤ 0.001; SGLT2I: Sodium-glucose cotransporter-2 inhibitors; DPP4I: Dipeptidyl peptidase-4 inhibitors; HR: hazard ratio; CI: confidence interval.

| | **New onset infective endocarditis HR[95% CI];P value** | **Cardiovascular mortality HR[95% CI];P value** | **All-cause mortality HR[95% CI];P value** |
|---|---|---|---|
| **SGLT2I v.s. DPP4I** | 0.51[0.38-0.68];<0.0001*** | 0.33[0.27-0.39];<0.0001*** | 0.29[0.23-0.36];<0.0001*** |

**Supplementary Table 6. Sensitivity analysis: Consideration of 1-year lag time effects in the SGLT2I v.s. DPP4I matched cohort.**

* for p≤ 0.05, ** for p ≤ 0.01, *** for p ≤ 0.001; SGLT2I: Sodium-glucose cotransporter-2 inhibitors; DPP4I: Dipeptidyl peptidase-4 inhibitors; HR: hazard ratio; CI: confidence interval.

| | **New onset infective endocarditis HR[95% CI];P value** | **Cardiovascular mortality HR[95% CI];P value** | **All-cause mortality HR[95% CI];P value** |
|---|---|---|---|
| **SGLT2I v.s. DPP4I** | 0.51[0.38-0.68];<0.0001*** | 0.29[0.24-0.36];<0.0001*** | 0.28[0.23-0.34];<0.0001*** |

**Supplementary Table 7. Sensitivity analysis: Risks of infective endocarditis upon excluding patients with drug abuse, developed infective endocarditis within 30 days after medications, patients with financial aid**

* for p≤ 0.05, ** for p ≤ 0.01, *** for p ≤ 0.001; SGLT2I: Sodium-glucose cotransporter-2 inhibitors; DPP4I: Dipeptidyl peptidase-4 inhibitors; HR: hazard ratio; CI: confidence interval.

| **SGLT2I v.s. DPP4I** | **New onset infective endocarditis**<br>**HR[95% CI];P value** |
|---|---|
| **Excluded patients with drug abuse** | 0.63[0.45-0.95];<0.0001*** |
| **Included patients who developed infective endocarditis within 30 days after medications** | 0.72[0.64-0.89];<0.0001*** |
| **Exclude patients with financial aid** | 0.62[0.35-0.78];<0.0001*** |

**Supplementary Table 8. Sensitivity analysis: Defining infective endocarditis by using two separate positive blood culture.**

| Infective endocarditis (blood culture only) | Number of patients | Number of events | Total person-year | Follow-up duration, years [95% CI] | Incidence [95% CI] per 100000 person-year | Adjusted hazard ratio [95% CI] | P-value |
|---|---|---|---|---|---|---|---|
| **DPP4I** | 28774 | 41 | 155050.4932 | 5.57 [5.26-5.8] | 26.43 [18.98-35.87] | 1 [reference] | NA |
| **SGLT2I** | 28774 | 22 | 159543.0877 | 5.62 [5.35-5.83] | 14.19 [9.92-21.48] | 0.57[0.34-0.95] | 0.0326* |
